# A topic landscape study for adolescent depression

**DOI:** 10.1101/2020.12.22.20248745

**Authors:** Tianran Li, Liang Guo, Xiaoqiang Wang, Stijn Decoster

## Abstract

Literature on adolescent depression is not only rich in content and diverse in form, but also scattered and complex in structure. However, there is no dynamic development analysis and evolution analysis related to the topic. Therefore, this study drew a topic landscape map and predicted the prevalence of topics in the future.

**Methods:** A systematic review was made to collect research publications on adolescent depression and describe the development of this field in the last five decades. We adopted visualization strategy and Herfindahl-Hirschman Index to uncover the latent structure of different topics from literature. The ARIMA model was utilized to predict the prevalence of each topic in the time dimension.

**Results:** By using content analysis technique, 21 topics are extracted from 1,595 articles of adolescent depression. These 21 topics were further divided into four hot topics, seven stable topics and ten cold topics based on the results of the ARIMA model. In particular, we predict that “family environment and parenting styles”, “diagnosis, treatment and interventions” and “mental disorder and behavior problems” will receive much attention in the next five years.

**Conclusion:** This paper provides temporal characteristics of each topic. This has an important implication to choose promising research topics for researchers and journal editors.

## Background

Over the years, the World Health Organization (WHO) has warned that depression is a major cause of disease and disability, affecting more than 264 million people worldwide. For example, depression is linked with a higher risk of suicide [1]. Studies have shown that depression increases significantly during adolescence and its course is frequently recurrent[1]. Depressive symptoms appearing in adolescence are related to more costly and frequent physical health problems[2], and increase the risk of suicide attempts[3].

Prior studies have investigated the explanatory theories of depression. Bernaras et al. (2019)[4] summarize a variety of causal factors, such as (1) biological factors including noradrenalin deficits, endocrine disorders, sleep-related disorders, alterations in brain structure, and genetics; (2) psychological factors including attachment theories, behavioral models, cognitive models, the self-control model, interpersonal theory, stressful life events, and sociocultural models. Extensive evidence illustrates that parenting styles or family situations are highly associated with adolescent depression[5, 6]. Parents with high conflicts, families with less warmth as well as aversiveness were found to augment the adolescent’s risk of depression. Parents also play an important role in prevention[7]. In addition, social networking sites can damage personal relationships and cause interpersonal-relationship conflicts. For instance, a mediation model was established to construct the relationship between social networking sites and adolescent depression [8].

Based on the classical theory of depression, a great number of studies have empirically explored the consequences as well as the determinants of adolescent depression. Multidisciplinary knowledge for adolescent depression is not only profound in content and diverse in form, but also scattered and complex in structure. To the best of our knowledge, there is no the topics of adolescent depression have been systematically reviewed across multiple disciplines in the past 50 years. There is also no a prediction of future prevalence of the topics in this literature is conducted. Hence, a clear mapping of this increasingly complex landscape is urgently established to help researchers make effective literature review. This extensive description of the field of adolescent depression will be valuable because it provides an understanding of how this field has evolved over time. Moreover, it reveals the consensus and differences among scholars, and reveals the blank of research on knowledge structure in this area.

In this study, we aim to predict the prevalence of topics that might generate groundbreaking research for adolescent depression in the future. A computerized overview of the literature in the past five decades is firstly carried out. We further uncover a latent structure which consisting of 21 topics and their development tracks. Finally, we predict the popularity of these 21 topics based on the temporal characteristics of each topic, which will help researchers and journal editors to choose promising research topics.

## Materials and Methods

### Data processing

Web of Science (WoS) core collection is the most authoritative citation index for scientific and scholarly research. We first collected the abstracts of all the articles that appeared in WoS. Notice that the first paper on adolescent depression appeared in 1967. Next, After removing duplicates, in total 2,072 articles were selected according to the following three criteria: 1) redaction in English; 2) including “adolescent depression” in title, abstract or keywords; 3) SCIE or SSCI indexed journals. After eliminating the articles which did not include an abstract and which were obviously unrelated to adolescent depression, we obtained a valid sample of 1,595 articles. The PRISMA flowchart in Appendix I illustrate the process of selecting or excluding the articles.

### Topic extraction methodology

We followed the conceptual analysis approach outlined by Hsieh & Shannon[9]. Firstly, we randomly split the sample articles into three sets: 600 articles in the first set as the “training sample”, 500 articles in the second set as the “validation sample” and the remaining 495 articles in the third set as the “test sample”. We invited 30 student volunteers who majored in social sciences to read the full text of the first set of articles. The students and the author team worked together to develop sets of terms and of coding rules. Next, the students and the author team developed a set of *n* topics based on these terms. Finally, each student and author read the abstracts of the 1,595 articles and assigned a topic loading (i.e. a score between zero to one that refers to how likely does a particular article belong to a particular topic) to every topic. If an article does not belong to a particular topic, then its topic loading to this topic is zero. We generated an article-topic matrix with a size of 1,595\**n*, in which each row represented an article and each column represented a topic. We assigned each article to the dominant topic with the highest topic loading.

In order to verify the effect of conceptual analysis and considering that there is no high linear correlation among the feature vectors, we employed the t-SNE approach to carry out nonlinear dimension reduction on the data[10]. A t-SNE algorithm can ensure that the distribution of data in low dimension has high similarity with the distribution of original feature space. Most importantly, it can visualize the distance between topics in two-dimensional space, so that we can have an intuitive idea about whether the classification of *n* topics makes sense or not.

Then, we followed the approach of Guo et al.[11]. adopted Herfindahl-Hirschman Index (HHI) to evaluate the level of topic diversity. HHI [12] is frequently used by economists and government regulators to measure the changes in market share, namely the dispersion degree of firm size in the market. Assuming that there are *N* enterprises in the industry, the value of HHI varies 1/*N* to 1. The higher the value, the higher the degree of market concentration and monopoly. HHI refers to the sum of squares of the percentage of total assets of each market competitor in an industry. Analogically, HHI in our case could be defined as the sum of squares of the ratio of the score of each article to the total score. The value of HHI for each topic further varies between 1/*N*_*i*_ and 1, where *N*_*i*_ is the number of articles with a score of not zero in topic *i*. However, the sample size of *N*_*i*_ is different for each topic. In order to make a fair horizontal comparison of concentration distribution between different topics, we normalized HHI for each topic as follows

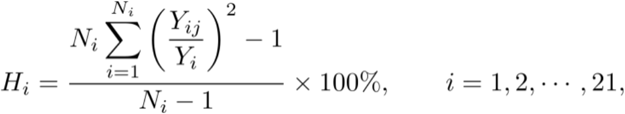

where *Y*_*ij*_ represents the score of article *j* in the topic *i* and *Y*_*i*_ the total score for the topic *i*. This normalized HHI takes value from zero to one. The larger the value of *H*_*i*_, the higher the degree of concentration of topic *i*.

### Topic dynamics methodology

We firstly classified each article according to the topic with the highest score, and counted the number of articles for each assigned topic. We further conducted time series analysis to predict the development trend of each topic in the next five years, and compared them with each other. The exponential smoothing model and the Autoregressive Integrated Moving Average (ARIMA) model are the two most widely used time series forecasting methods. Based on these two prediction methods, many other prediction methods have been born. In our case, most of the article count series of 21 topics are observed to involve autocorrelation. This fact will lead to the correlated residuals when fitting data by the regression model. Obviously, the phenomenon of autocorrelation hinder the performance of regression model to fit our data. Such defect of autocorrelation can be solved by the I (“integrated”) part in ARIMA. The article count sequence values will be differentiated between the current data and the previous one or several data, so that non-stationary series can be modeled. In the ARIMA model, the AR part is responsible for the linear regression of the previous time series value; the MA part is responsible for the linear regression of the previous random error series. ARIMA clearly meets a set of standard structures in time series data (i.e. article count series data) and provides a simple and powerful method for proficient time series forecasting [13].

According to *Time Series Analysis Forecasting and control* [14], the timing data predicted by the ARIMA model must be stable, which, by definition, has no trend and no periodicity (that is, its mean value has a constant amplitude on the time axis, and its variance tends to the same stable value on the time axis). We used Dickey-Fuller Test to judge the stability of time series data, which is a common unit root Test method. For the time series data that do not meet the requirements of stationarity, we made a difference to eliminate the periodic factors. The degree of differences to make the data stable was taken as parameter *d*. If the preprocessed time series was determined to be a stable non-white noise sequence, we determined the autocorrelation function (ACF) and partial autocorrelation function (PACF) as parameters *p* and *q*. We conducted a grid-search to calculate the scores of RMSE, AIC and BIC under different combinations of orders, and chose the order specifications with the lowest scores. To ensure the validity of the model, we drew the residual to check if it is completely random white noise, and performed the Ljung-Box test to formally check whether the error is uncorrelated among many lags [15]. Otherwise, we will improve the model by removing all the remaining trends.

## Results

### Data description

The top line in Figure 1 depicts the annual growth rate of article count from 1967 to 2020.

**Figure 1.**
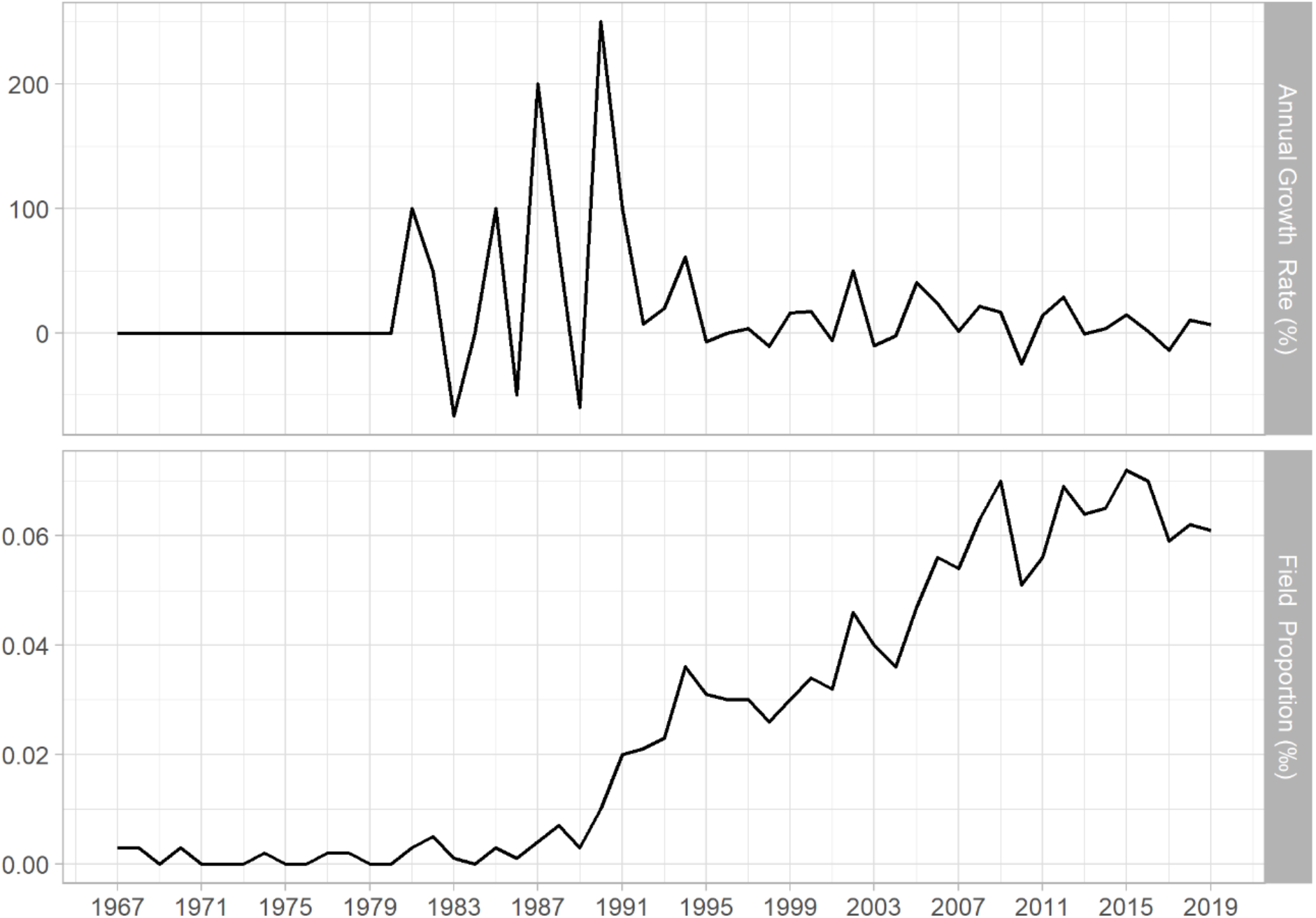
The percentage of publications (‰, top) and the percentage of the growth rate (%, bottom) in the field of “adolescent depression”.

It can be seen that after a period of volatility from 1980 to 1995, the growth rate of papers has tended to be flat in the past two decades. In 1987, the growth rate jumped from −50.00% of the previous year to 200.00%, and reached the highest level of 250.00% in 1990. However, as shown by the bottom line in Figure 1, the proportion of the articles in the field of adolescent depression to the total SCIE or SSCI indexed articles has become larger and larger since 1990. It means that over the past five decades, the field has been greatly developed. Especially, only one article (0.003‰) related to adolescent depression is retrieved in 1967 in WoS, but this number has grown to 140 (0.061‰) in 2019. The average annual growth rate in this field reached 18.7%.

The papers related to the adolescent depression were found to be published in 586 journals. The authors of these papers are distributed in 71 countries, of which the United States accounts for 61.15%, Britain 9.56%, Canada 7.42%, Australia 6.95%, and China 4.54%. One twentieth of them come from the University of Pittsburgh. Table 1 summarizes the ten most frequent organizations, ten most frequent fields and ten most frequent outlets. We notice that the top ten fields listed in Table 1 account for about 92.13% of all articles. Additionally, the articles are spread across 79 WoS research fields.

**Table 1.**
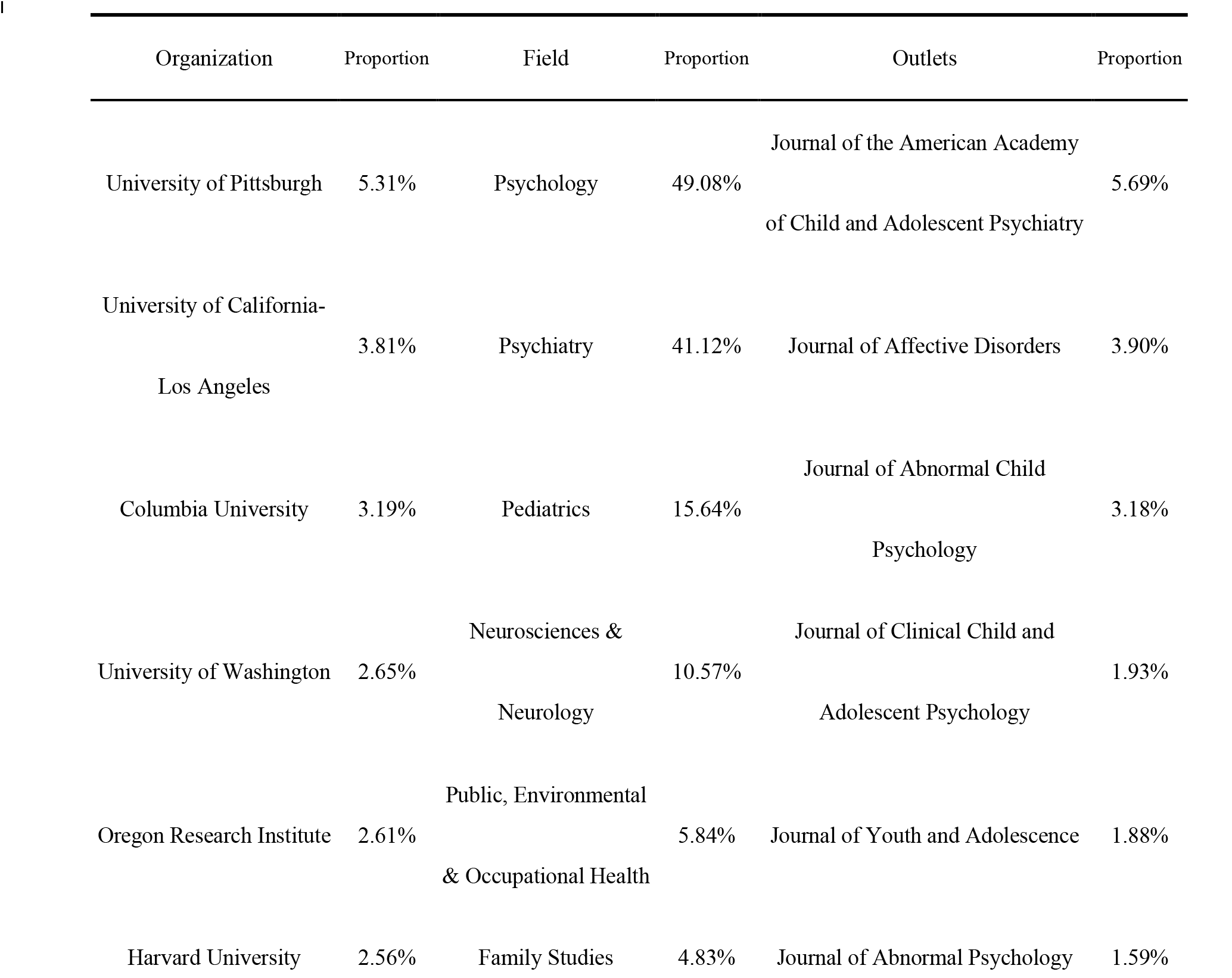
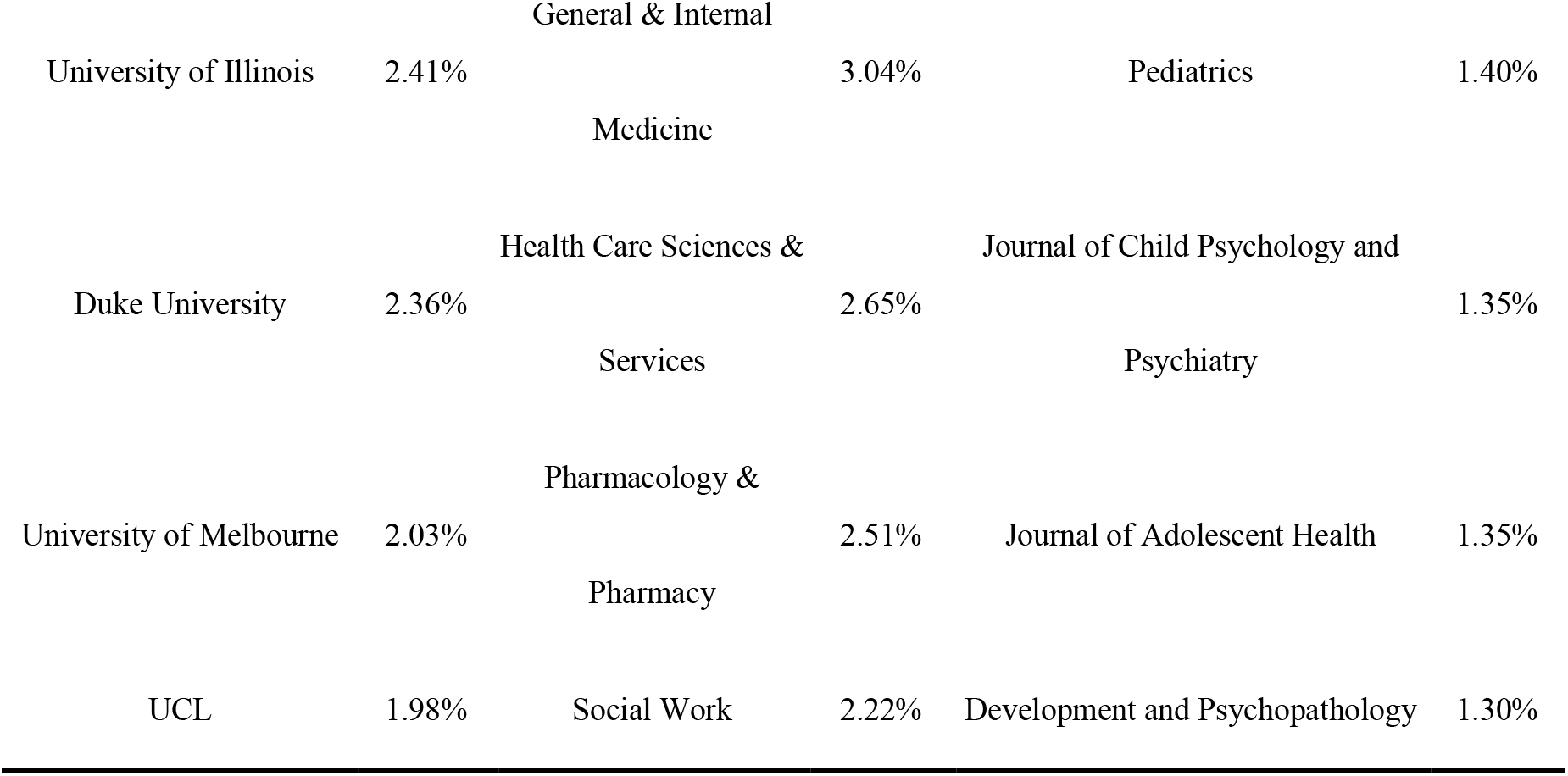
Top 10 organizations, research fields and research outlets.

### Topic extraction

Following the method described in Subsection *Topic extraction methodology*, the students and the author team initially classified the second set of 500 articles into 20 topics. However, we found that the optimal number of topics should be 21. Finally, the students and the author team classified the third set of articles into 21 topics (see Table 2). We believe that these 21 topics allowed to correctly categorize all the sample articles. In Figure 2, we find that the topics are evenly distributed, which validate our conceptual analysis and topic distribution.

**Table 2.**
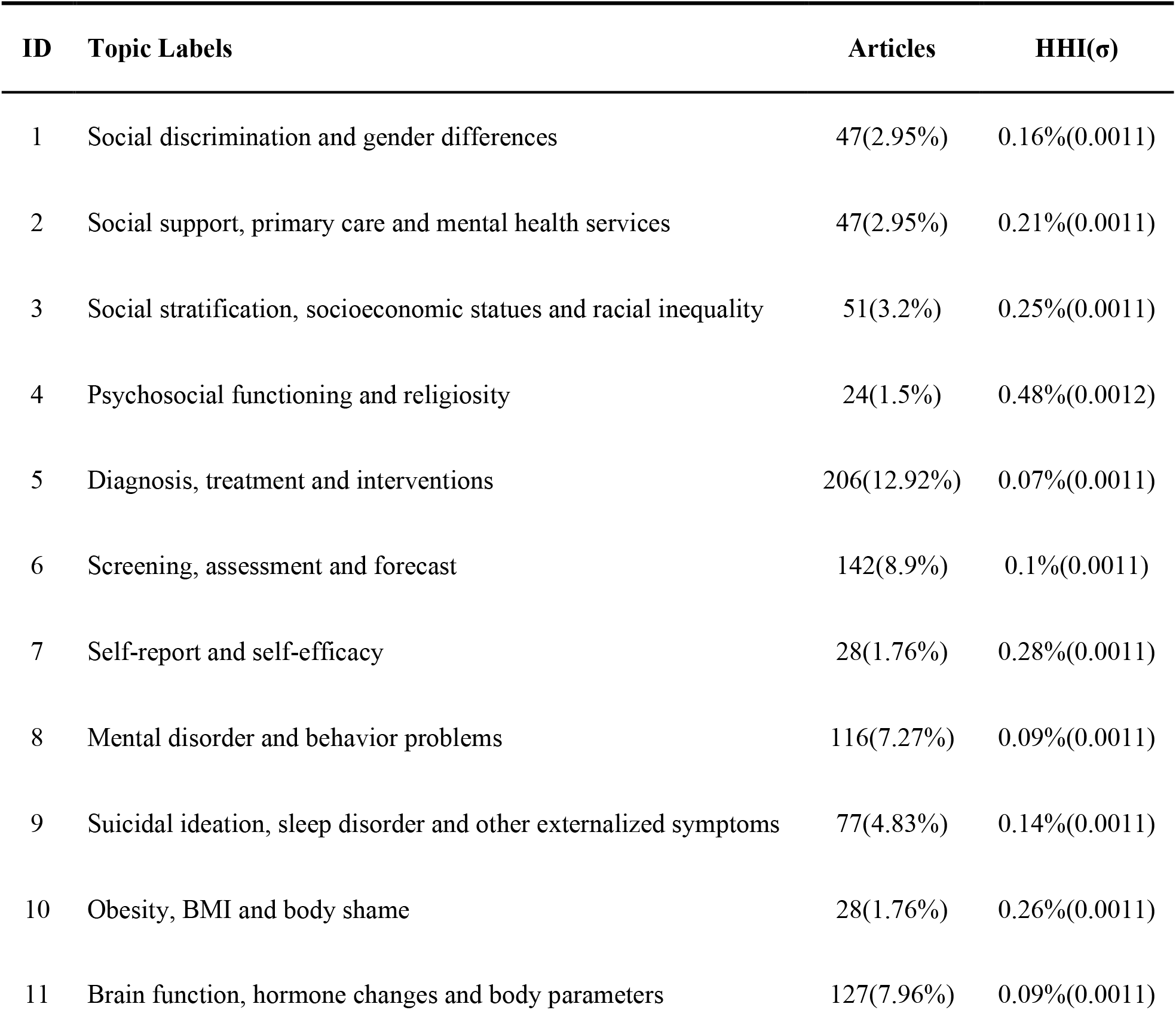
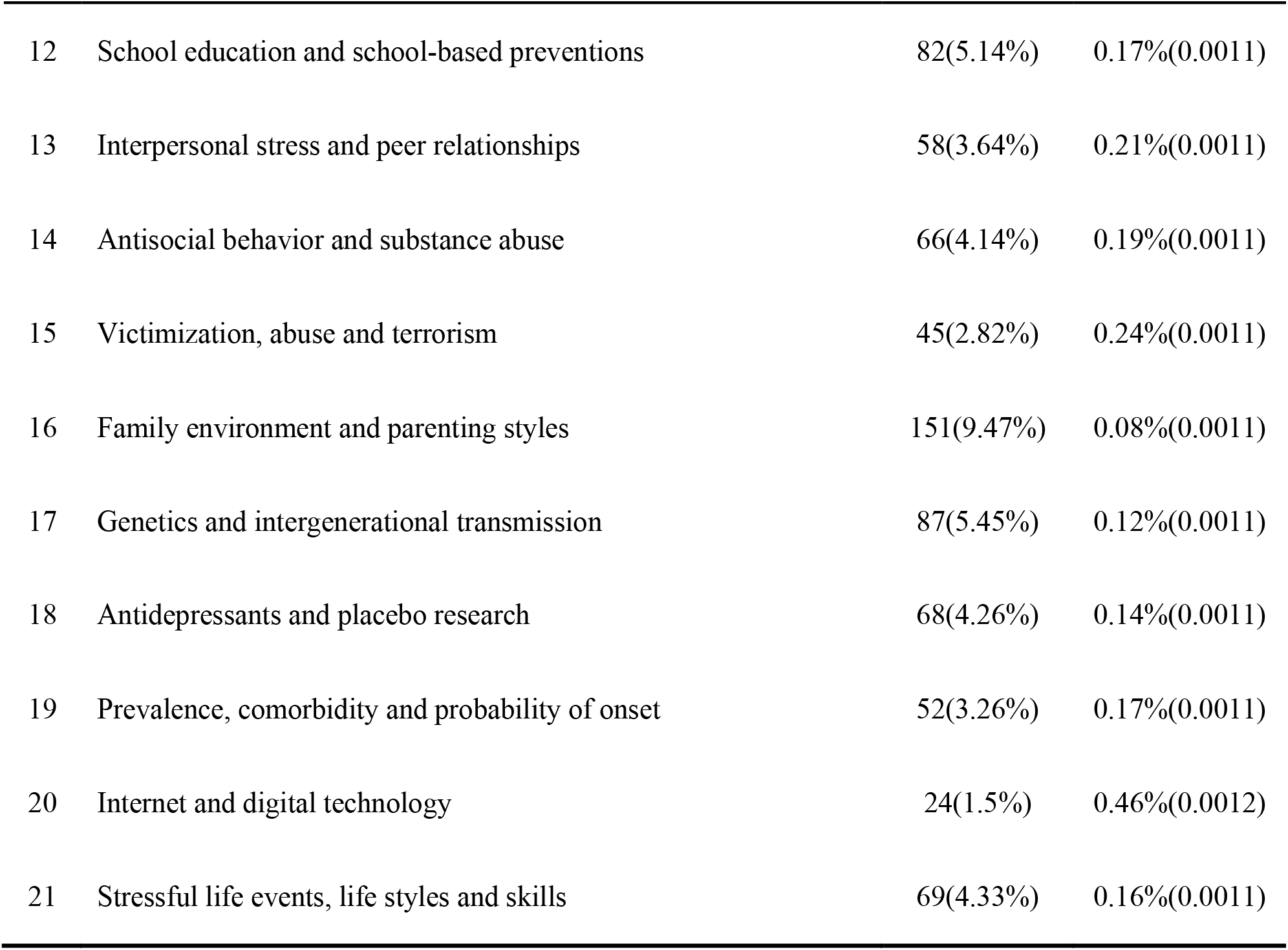
The 21 topics and their Herfindahl-Hirschman Index (HHI)

| ID | Topic Labels | Articles | HHI($\sigma$) |
| --- | --- | --- | --- |
| 1 | Social discrimination and gender differences | 47(2.95%) | 0.16%(0.0011) |
| 2 | Social support, primary care and mental health services | 47(2.95%) | 0.21%(0.0011) |
| 3 | Social stratification, socioeconomic statues and racial inequality | 51(3.2%) | 0.25%(0.0011) |
| 4 | Psychosocial functioning and religiosity | 24(1.5%) | 0.48%(0.0012) |
| 5 | Diagnosis, treatment and interventions | 206(12.92%) | 0.07%(0.0011) |
| 6 | Screening, assessment and forecast | 142(8.9%) | 0.1%(0.0011) |
| 7 | Self-report and self-efficacy | 28(1.76%) | 0.28%(0.0011) |
| 8 | Mental disorder and behavior problems | 116(7.27%) | 0.09%(0.0011) |
| 9 | Suicidal ideation, sleep disorder and other externalized symptoms | 77(4.83%) | 0.14%(0.0011) |
| 10 | Obesity, BMI and body shame | 28(1.76%) | 0.26%(0.0011) |
| 11 | Brain function, hormone changes and body parameters | 127(7.96%) | 0.09%(0.0011) |
| 12 | School education and school-based preventions | 82(5.14%) | 0.17%(0.0011) |
| 13 | Interpersonal stress and peer relationships | 58(3.64%) | 0.21%(0.0011) |
| 14 | Antisocial behavior and substance abuse | 66(4.14%) | 0.19%(0.0011) |
| 15 | Victimization, abuse and terrorism | 45(2.82%) | 0.24%(0.0011) |
| 16 | Family environment and parenting styles | 151(9.47%) | 0.08%(0.0011) |
| 17 | Genetics and intergenerational transmission | 87(5.45%) | 0.12%(0.0011) |
| 18 | Antidepressants and placebo research | 68(4.26%) | 0.14%(0.0011) |
| 19 | Prevalence, comorbidity and probability of onset | 52(3.26%) | 0.17%(0.0011) |
| 20 | Internet and digital technology | 24(1.5%) | 0.46%(0.0012) |
| 21 | Stressful life events, life styles and skills | 69(4.33%) | 0.16%(0.0011) |

**Figure 2.**
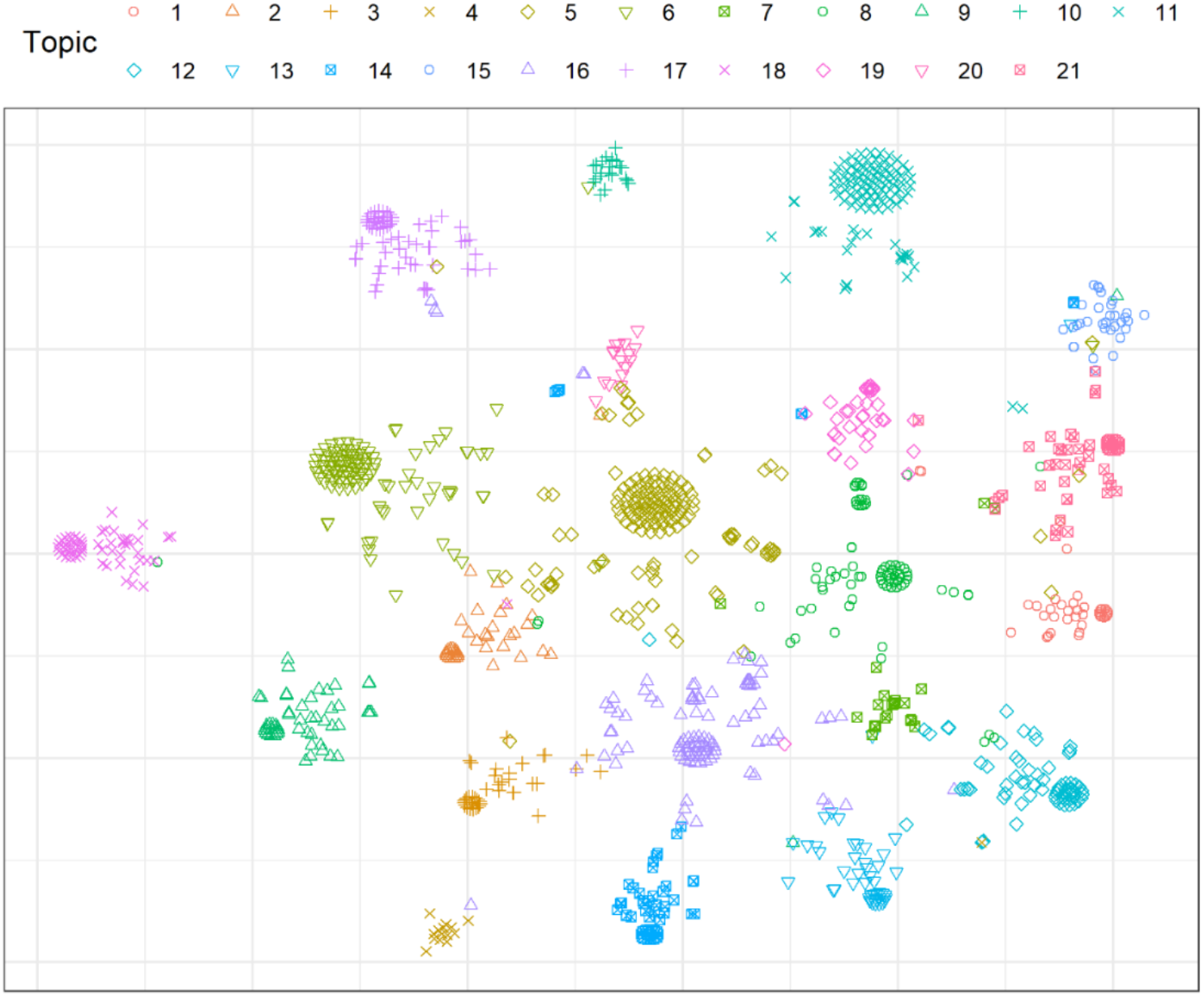
t-SNE clustering of the 1,595 articles in two-dimensional maps. The analysis was statistically constrained to 21 clusters for distinct topics as colored nodes.

Table 2 lists the score of normalized HHI for all the 21 topics. Some topics exhibit a high degree of concentration. For example, “psychosocial functioning and religiosity” (Topic 4), “internet and digital technology” (Topic 20) and “self-report and self-efficacy” (Topic 7) exhibit the three highest HHI, and they are all topics with strong topicality, resulting in a relatively concentrated distribution of scores.

Topics exhibiting the lowest HHI include “diagnosis, treatment and interventions” (Topic 5), “family environment and parenting styles” (Topic 16) and “brain function, hormone changes and body parameters” (Topic 11) discussed “treatment” and “interventions”, “family environment” and “physiological factors”, etc., which are easy to be mentioned incidentally, so the score distribution is relatively uniform.

Notice that we obtained consistent results between t-SNE clustering and degree of concentration (HHI in Table 2). For instance, the lower HHI scores are more scattered in the topic area shown in Figure 2 (e.g. Topic 5, 6, 11, 16). Conversely, the higher HHI score shows a larger cluster in the topic area shown in Figure 2 (e.g. Topic 4, 7, 20). These results confirm the rationality of our selected topics.

### Topic dynamics

Table 2 displays the article count data in each topic. The article count for each topic ranges from 24 to 206 with mean 75.95 and standard deviation 47.77. “diagnosis, treatment and interventions” (Topic 5), “family environment and parenting styles” (Topic 16) and “screening, assessment and forecast” (Topic 6) are the three most prevalent topics. In contrast, “self-report and self-efficacy” (Topic 7), “psychosocial functioning and religiosity” (Topic 4), and “Internet and digital technology” (Topic 20) are the three least prevalent topics.

In order to study the figures and publications that have contributed the most to each topic in the field of “adolescent depression”, we listed the top authors and publications that appear the most frequently on each topic in AppendixII, which can provide reference for research direction and popular trends in this field. Note that we listed multiple authors or publications if their frequencies of appearance were the same. We found that Peter M. Lewinsohn from Oregon Research Institute was one of the most productive and influential authors in Topic 1, 4, and 19. We also found that the Journal of The American Academy of Child and Adolescent Psychiatry was the most popular outlet for nine topics. Finally, we selected three most representative articles (i.e. with the highest topic loadings) of each topic and summarize the information in AppendixII.

Notice that the number of articles published in the field of adolescent depression increased from one in 1967 to 140 in 2019, which indicate that more and more scholars have participated and made contributions in this field. Further, we expected to explore the rapid development period and the dynamic development law for each topic in the time dimension. In order to show the process more intuitively, we drew the ThemeRiver (see Appendix III), which indicated that Topic 2 (social support, primary care and mental health services), Topic 5 (diagnosis, treatment and interventions), Topic 6 (screening, assessment and forecast), Topic 11 (brain function, hormone changes and body parameters), and Topic 16 (family environment and parenting styles) experienced rapid growths during the last decade.

Then, we built 22 time series comprising of the field integrally and 21 topics. An optimized ARIMA model is finally adopted to predict the proportion of publications in this field for each topic in the next five years (i.e. 2021–2025), respectively. The predicted average annual growth rate will be used as the indicator of the prevalence of future topic (see Table 3). In the future, the field is likely to expand steadily because its annual growth rate will reach 49.03%, but this figure is far lower than the peak annual growth rate of 250% in 1990. This fact indicates that adolescent depression will continue to be a hotspot of academic research, but it is unlikely to attract explosive attention in professional research communities.

**Table 3.** The results of ARIMA and prediction.

| Topic | Order | logLik | AIC | BIC | HQIC | RMSE | LBstat | Category |
| --- | --- | --- | --- | --- | --- | --- | --- | --- |
| The Field | (0,2,1) | -124.85 | 257.7 | 264.92 | 260.39 | 18.61 | 0.54 | Benchmark(49.03%) |
| Topic 1 | (0,1,1) | -40.82 | 87.64 | 91.17 | 88.58 | 1.3 | 0.21 | Cold(-35.58%) |
| Topic 2 | (0,1,0) | -15.23 | 36.46 | 39.73 | 37.23 | 1.05 | 0.87 | Stable(22.34%) |
| Topic 3 | (4,1,2) | -29.58 | 73.16 | 79.77 | 74.28 | 1.11 | 0.83 | Cold(-131.1%) |
| Topic 4 | (0,1,0) | -4.72 | 15.44 | 17.76 | 15.56 | 0.8 | 0.57 | Cold(-12.35%) |
| Topic 5 | (0,1,1) | -82.33 | 166.81 | 171.3 | 168.32 | 2.69 | 0.27 | Hot(98.75%) |
| Topic 6 | (2,1,0) | -77.86 | 159.62 | 167.54 | 162.38 | 1.89 | 0.85 | Stable(1.26%) |
| Topic 7 | (0,2,1) | -18.02 | 44.03 | 46.86 | 44 | 0.91 | 0.72 | Stable(33.54%) |
| Topic 8 | (0,1,1) | -67.07 | 136.91 | 140.91 | 138.14 | 2.43 | 0.13 | Hot(82.47%) |
| Topic 9 | (0,1,1) | -51.68 | 108.94 | 112.93 | 110.16 | 1.52 | 0.33 | Cold(-46.41%) |
| Topic 10 | (0,2,2) | -26.71 | 61.43 | 63.99 | 61.2 | 1.46 | 0.71 | Cold(-35.79%) |
| Topic 11 | (1,0,0) | -64.76 | 135.52 | 139.29 | 136.61 | 2.91 | 0.74 | Hot(68.24%) |
| Topic 12 | (0,2,1) | -53.36 | 103.64 | 108.01 | 104.67 | 1.87 | 0.71 | Cold(-131.74%) |
| Topic 13 | (0,1,1) | -35.98 | 77.97 | 81.1 | 78.65 | 1.33 | 0.73 | Cold(-7.34%) |
| Topic 14 | (0,1,1) | -38.28 | 82.57 | 85.7 | 83.25 | 1.47 | 0.45 | Cold(-1.77%) |
| Topic 15 | (0,1,1) | -31.26 | 68.53 | 70.85 | 68.65 | 1.57 | 0.39 | Stable(30.25%) |
| Topic 16 | (1,1,1) | -73.99 | 155.97 | 161.71 | 157.84 | 2.56 | 0.94 | Hot(106.2%) |
| Topic 17 | (0,1,3) | -50.6 | 107.21 | 110.86 | 108.22 | 1.8 | 0.62 | Stable(5.65%) |
| Topic 18 | (0,1,1) | -49.21 | 104.42 | 108.2 | 105.51 | 1.59 | 0.46 | Cold(-6.74%) |
| Topic 19 | (0,0,0) | -38.54 | 81.08 | 83.35 | 81.65 | 1.29 | 0.38 | Stable(45.22%) |
| Topic 20 | (0,0,2) | -23.43 | 50.86 | 51.66 | 50.36 | 2.04 | 0.23 | Stable(43.64%) |
| Topic 21 | (0,1,0) | -13.41 | 32.83 | 36.24 | 33.69 | 1.35 | 0.45 | Cold(-45.93%) |

We utilize the following criteria to divide 21 topics into three categories: hot topics with predicted annual growth rate greater than or equal to the field growth rate (i.e. 49.03%); stable topics with growth rate greater than zero or equal to zero but less than the field growth rate; cold topics with negative topic rate. As a result, we find four hot topics, seven stable topics and ten cold topics (see Table 3 and Table 2).

## Discussion

Our ARIMA model predicted that “School education and school-based preventions” (Topic 12, - 131.74%), “Social stratification, socioeconomic statues and racial inequality” (Topic 3, −131.1%) and “Suicidal ideation, sleep disorders and other externalized symptoms” (Topic 9, −46.41%) might dramatically shrink. Likewise, “Obesity, BMI and body shame” (Topic 10, −35.79%) and “Psychosocial functioning and religiosity” (Topic 4, −12.35%) also showed a significant downward trend. That is probably because these topics have been addressed extensively over decades, which makes these fields of investigation saturated.

On the contrary, the “Family environment and parenting styles” (Topic 16, 106.2%) and “Diagnosis, treatment and interventions” (Topic 5, 98.75%) showed a significantly positive trend of increase. In the past studies in this field, these two topics also showed a high degree of popularity (22.38% of the articles). Our ARIMA findings suggested that given the omnipresence of work-life imbalance in today’s industrial relations, it is reasonable that the research focus of the field might turn to family environment and treatment diagnosis. In addition, “Brain function, hormone changes and body parameters” (Topic 11, 68.24%) and “Prevalence, co-morbidity and probability of onset” (Topic 19,45.22%) also presented a sign of popularity. This might suggest that more and more researchers will develop and employ new scientific methods.

It is worth mentioning that although “Internet and digital technology” (Topic 20, 43.64%) was a small topic in the past, our ARIMA analysis predicted that this topic might attract attention in the future, probably because on the one hand, more and more adolescents are addicted to the Internet, which may lead to depression; on the other hand, future research may investigate how to use the Web and digital technology to prevent adolescent depression.

This study constructed all of the WoS literature in the field of adolescent depression over the last five decades. By describing the entire knowledge system at a relatively detailed level, we contribute to a rich understanding of the topic landscape of the field so that researchers and journal editors can appreciate the entire range of topics and choose the topics they wish to study in depth. In addition, because we help instructors identify important topics and assign groundbreaking articles related to each topic, the topic landscape provides information for related teaching and curriculum design. However, it must be pointed out that we only collected sample articles from WoS. Although WoS is the most comprehensive and authoritative source for high-impact publications, we may run the risk of missing articles published in emerging journals or books. Consequently, our findings may be biased. Future studies may also collect publication records from other freely-accessible online publications.

## Conclusions

We firstly established a systematic overview of the research on adolescent depression in the last five decades: its evolution, topic landscape, and dynamics. We uncovered the latent structure of 21 different topics from 1,595 articles related to adolescent depression. Secondly, we utilized the ARIMA model to predict the prevalence of 21 topics in the next five years. Our forecasting results suggested that although this field might continue to be a hot research direction and attract more attention, ten topics might lose their popularity.

## Data Availability

Some or all data, models, or code generated or used during the study are available from the corresponding author by request.

## Ethics and consent

N/A

## Appendix I

**Figure.**
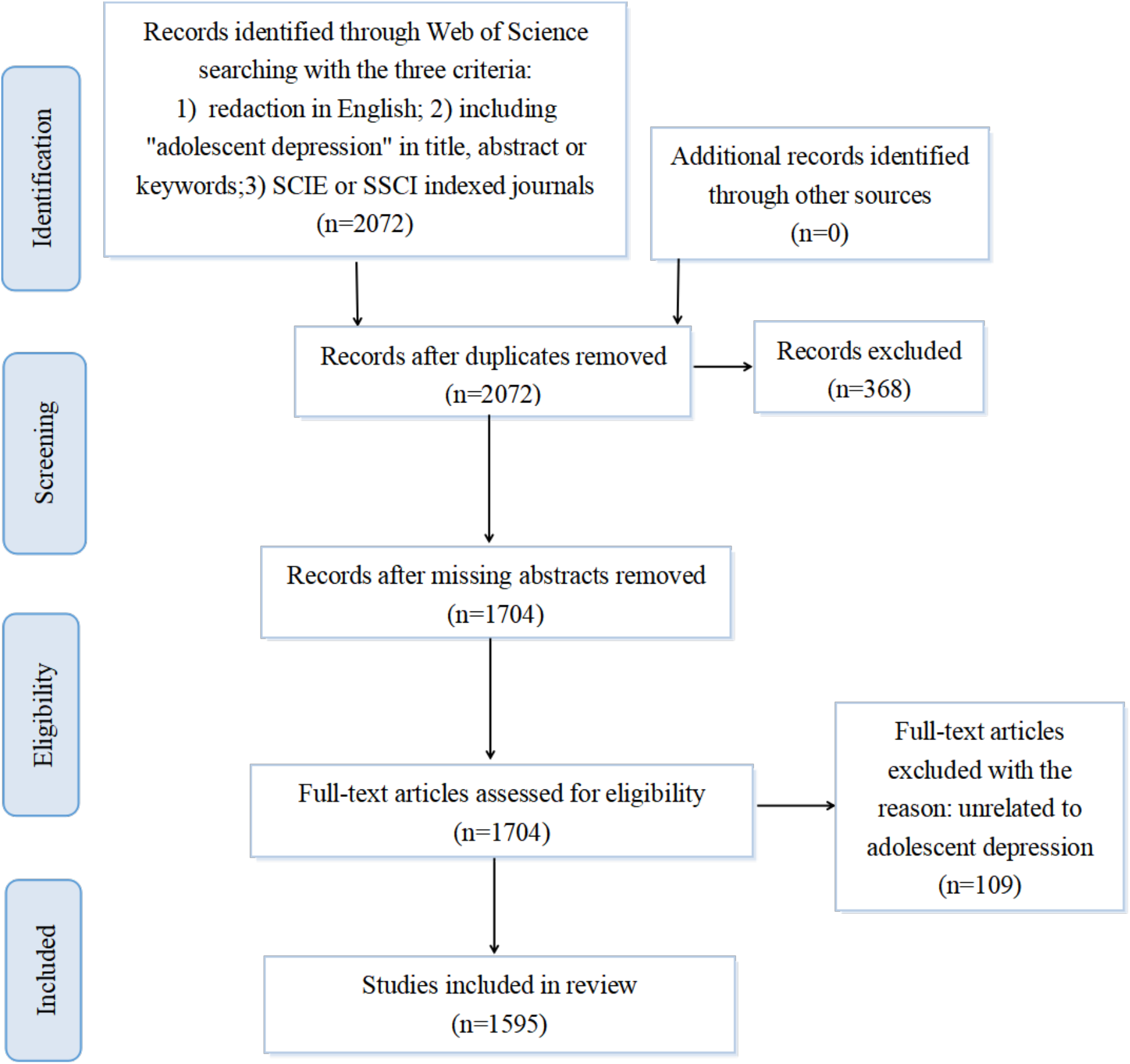
PRISMA flowchart of literature search and identification.

## Appendix II

**Table 1.** The most popular authors,outlets and the three most representative articles per topic.

| Topic | Authors | Outlets | Representative articles |
| --- | --- | --- | --- |
| Topic 1 | Lewinsohn, PM ;<br>Hops, H | Journal of Abnormal Psychology | Avison & McAlpine, 1992 [1];<br>Cyranowski, Frank, Young, & Shear,<br>2000[2]; Han, 2014[3]. |
| Topic 2 | Garralda, E | Journal of Adolescent Health ;<br>Journal of Affective Disorders | Draucker, 2005[4]; Jacobson, 1991[5];<br>Stein, Zitner, & Jensen, 2006[6]. |
| Topic 3 | Wickrama, K. A.<br>S. | Journal of Adolescent Health | Aslund, Leppert, Starrin, & Nilsson,<br>2009[7]; Respress, Morris, Gary, Lewin,<br>& Francis, 2013[8]; Su, 2010[9]. |
| Topic 4 | Lewinsohn, PM ;<br>Seeley, JR | Psychological Medicine ; Journal of<br>Abnormal Child Psychology ;<br>Journal of The American Academy of<br>Child and Adolescent Psychiatry | Derdikman-Eiron et al., 2011[10];<br>Purwono & French, 2016[11]; Rose, Rose,<br>Miller, & Dyer, 2018[12]. |
| Topic 5 | Birmaher, B | Journal of The American Academy of<br>Child and Adolescent Psychiatry | Fletcher, 2008[13]; Garland, 1994[14];<br>Kaslow & Thompson, 1998[15]. |
| Topic 6 | McCauley,<br>Elizabeth ;<br>Birmaher, B | Journal of The American Academy of<br>Child and Adolescent Psychiatry | Benavidez & Matson, 1993[16]; Gray,<br>Dubin-Rhodin, Weller, & Weller,<br>2009[17]; LeBlanc, Almudevar, Brooks, &<br>Kutcher, 2002[18]. |
| Topic 7 | Orchard, Faith ;<br>Reynolds, Shirley | Journal of Child And Family<br>Studies ; Journal of Affective<br>Disorders | Joshi & Mehta, 2017[19]; Manikam,<br>Matson, Coe, & Hillman, 1995[20];<br>Minnix, Blalock, Marani, Prokhorov, &<br>Cinciripini, 2011[21]. |
| Topic 8 | Collishaw,<br>Stephan | Journal of The American Academy of<br>Child and Adolescent Psychiatry | Bardone et al., 1998[22]; Benamos,<br>1992[23]; JacksonWalker & Nitz,<br>1996[24]. |
| Topic 9 | Rao, U ; DAHL,<br>RE | Journal of The American Academy of<br>Child and Adolescent Psychiatry | Becker, Langberg, & Evans, 2015[25];<br>Sanchis & Simon, 2012[26]; Spirito,<br>Francis, Overholser, & Frank, 1996[27]. |
| Topic 10 | Richardson, Laura<br>P. | General Hospital Psychiatry | Roberts & Duong, 2015[28]; Wardle,<br>Williamson, Johnson, & Edwards,<br>2006[29]; You, Shin, & Kim, 2017[30]. |
| Topic 11 | Yang, Tony T. | Journal of Affective Disorders | Karabel et al., 2016[31]; Merz, He, Noble,<br>& Pediatric Imaging Neurocognition,<br>2018[32]; Sokolov, Kutcher, & Joffe,<br>1996[33]. |
| Topic 12 | Beaudry, Mary Beth ; Swartz, Karen ; Wilcox, Holly | School Mental Health | Possel, Horn, & Hautzinger, 2006[34]; Ruble, Leon, Gilley-Hensley, Hess, & Swartz, 2013[35]; Shirk, Kaplinski, & Gudmundsen, 2009[36]. |
| Topic 13 | Mufson, Laura | Journal of Abnormal Child Psychology | Auerbach, Ho, & Kim, 2014[37]; Hammen, 2009[38]; La Greca & Harrison, 2005[39]. |
| Topic 14 | Cullen, W. | Journal of Youth And Adolescence ; Journal of The American Academy of Child and Adolescent Psychiatry | Deykin, Levy, & Wells, 1987[40]; Harrington, Fudge, Rutter, Pickles, & Hill, 1991[41]; Subramaniam, Harrell, Huntley, & Tracy, 2009[42]. |
| Topic 15 | Shahar, Golan | Child Abuse & Neglect | Davis et al., 2019[43]; Henrich & Shahar, 2008[44]; Resett & Caino, 2019[45]. |
| Topic 16 | Jorm, Anthony F. ; Allen, Nicholas B. | Journal of The American Academy of Child and Adolescent Psychiatry | Cardamone-Breen, Jorm, Lawrence, Mackinnon, & Yap, 2017[46]; Fard, Gharraee, Farid, & Aghebati, 2017[47]; Shiner & Marmorstein, 1998[48]. |
| Topic 17 | Hammen, Constance ; Thapar, Anita | Journal of The American Academy of Child and Adolescent Psychiatry | Eley et al., 2004[49]; Lee & Koeske, 2010[50]; Rice, 2010[51]. |
| Topic 18 | Bylund, David B. | Journal of The American Academy of Child and Adolescent Psychiatry | Emslie et al., 2008[52]; Evans, Ha, Saisch, Ellison, & Fombonne, 1998[53]; Kutcher et al., 1994[54]. |
| Topic 19 | Lewinsohn, Peter M. ; Seeley, John R. | Journal of Affective Disorders | Hammen, Brennan, Keenan-Miller, & Herr, 2008[55]; Quijada et al., 2005[56]; Rey, 1994[57]. |
| Topic 20 | Abeles, Paul ; Griffiths, Kathleen M. ; Lei, Li | Behavioural and Cognitive Psychotherapy ; Computers In Human Behavior ; Jmir Research Protocols | Ha et al., 2007[58]; Lindqvist et al., 2020[59]; Ophir, Asterhan, & Schwarz, 2019[60]. |
| Topic 21 | Harkness, Kate L. | Journal of Abnormal Psychology | Boardman, Alexander, & Stallings, 2011[61]; Lessem et al., 1996[62]; Lopez, Fuentes, Palos, & Oudhof, 2012[63]. |

## Appendix III

**Figure.**
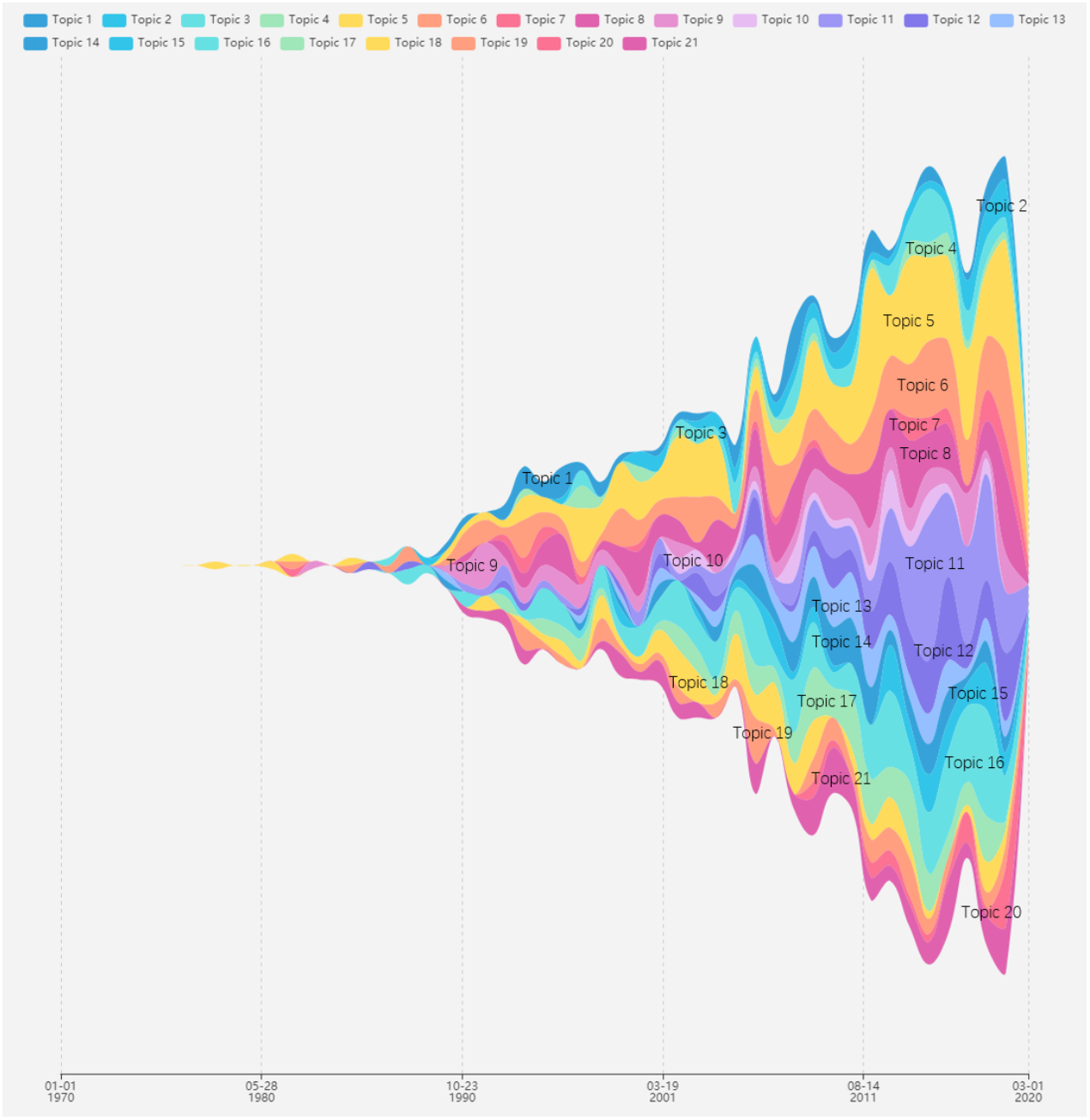
ThemeRiver drawing for topics developmen.

